# An exploratory investigation of placental metabolomic alterations associated with maternal smoking

**DOI:** 10.64898/2026.02.19.26346613

**Authors:** Wadzanai Masvosva, Retu Haikonen, Teemu Gunnar, Marko Lehtonen, Leea Keski-Nisula, Jaana Rysä, Olli Kärkkäinen

**Affiliations:** School of Pharmacy, University of Eastern Finland, Kuopio, Finland; Institute of Public Health and Clinical Nutrition, University of Eastern Finland, Kuopio, Finland; Forensic Chemistry, Department of Safety, Finnish Institute for Health and Welfare, Helsinki, Finland; Institute of Clinical Medicine, School of Medicine, University of Eastern Finland, Kuopio, Finland; Department of Obstetrics and Gynecology, Kuopio University Hospital, Kuopio, Finland

**Keywords:** tobacco, smoking, nicotine, metabolomics, placenta

## Abstract

Maternal smoking during pregnancy is associated with adverse effects on offspring health through impaired placental structure and function. Nicotine and other tobacco-related compounds readily cross the placental barrier, disrupt metabolic pathways, and increase the risk of long-term developmental disorders in newborn. Here, placental metabolic alterations associated with maternal smoking exposure were examined with metabolomics. We used placental samples from the Kuopio Birth Cohort study from 23 nonsmoking controls pregnancies, 19 pregnancies with early smoking exposure (cotinine detected in first-trimester but not in at-term samples), and 13 pregnancies with continuous smoking-exposure (cotinine detected in both first-trimester and at-term samples). Differences in placental metabolomic profiles were seen between controls and both smoking-exposed groups. For example, increased activity of xenobiotic metabolism pathways showed as elevated CYP1A2-related metabolites, e.g., aminoamide local anesthetic metabolite detected in both smoking-exposure groups (p=0.0042 and 0.0019, respectively). Disruptions in amino acid metabolism were observed, e.g., reduced placental tryptophan levels (p=0.0209 and 0.0237). Placentas from women who quit smoking during showed markers of reduced oxidative stress, lower oxidized glutathione (p=0.0119) and higher ergothioneine (p=0.0426) levels. These findings indicate that many smoking-related effects on the placental metabolome persist beyond acute nicotine exposure, showing long-term biological effects of maternal smoking during pregnancy.

**Plain language summary:** Smoking during pregnancy can possibly change how the placenta functions, which also affects the newborn’s long-term health. In this study, we compared placentas from nonsmokers, women who quit during pregnancy, and those who kept smoking. Clear chemical differences were seen in the placentas of smoking exposed pregnant women. The main changes included lowered levels of tryptophan and glutathione, which are important for growth and protection from stress. These results show that smoking-related changes in the placenta can persist beyond active nicotine exposure.

## Introduction

Smoking during pregnancy remains one of the leading preventable risk factors for adverse pregnancy outcomes worldwide^1^. The effects of maternal smoking during the pregnancy may have permanent effects to newborn which may last to adulthood^2^. A retrospective cohort study demonstrated that maternal cigarette consumption as low as 1-5 cigarettes per day before and during pregnancy, may increase the risk of birth congenital anomalies including limb reduction defect, cleft lip and hypospadias^3^. Such exposure has also been linked to neurological, cognitive, cardiovascular and respiratory impairments in offspring^4–6^.

There are benefits for both the mother and fetus if smoking cessation happens before the 15^th^ gestational week^7^. Pregnant women who have difficulty quitting smoking are more likely to succeed after receiving nicotine replacement therapy^8^. Furthermore, in nonpregnant populations, electronic cigarettes better known as vaping have been marketed as a safer and healthier smoking alternative to encourage smoking cessation^9^.

However, evidence shows that nicotine in itself is harmful during pregnancy, and that electronic cigarettes can induce similar effects as smoking on lung function and cardiovascular function^10^. Considering this, it has been established that there are no safe levels nor a safe trimester for maternal exposure to tobacco or nicotine in pregnancy^11^. Pregnancy increases nicotine metabolism due to enhanced activity of the cytochrome P450 (CYP) enzyme CYP2A6, resulting in higher nicotine clearance rates^12^. Consequently, many pregnant smokers must consume greater amounts of nicotine to achieve the desired effect, thereby increasing fetal exposure^13^.

Nicotine is able to cross the blood-placental barrier and may accumulate in the fetal circulation^14^. It adversely affects fetal tissue development and function, and is associated with long-term metabolic disturbances in the offspring^15^. One of the principal detrimental effects of smoking is the reduced placental blood flow, which limits the delivery of oxygen and nutrients to the developing fetus^16^. Proper placental development and function are important for a healthy pregnancy and fetus development^14^. Studies on the effects of smoking on the placenta have shown that it interferes with placentation, thereby reducing the placenta’s capacity to establish and maintain an optimal supply of nutrients and oxygen to the fetus^17^. Moreover, smoking has been linked to both increased and reduced placental weight, underlying a complex impact on placental physiology^18,19^. Epigenetic modifications, such as an altered DNA methylation in growth-related genes, have been reported in the placental tissue of smoking mothers and are believed to exert long-term effects on offspring growth^20^. Smoking also disrupts placental enzyme systems involved in hormone metabolism, including those regulating progesterone, a hormone critical for maintaining pregnancy and supporting fetal neuroendocrine development^21^. Maternal smoking promotes oxidative stress and inflammation within placental tissue, contributing to endothelial dysfunction and cellular injury^22^. Elevated production of reactive oxygen species (ROS) may deplete antioxidant defenses, such as glutathione, potentially impairing fetal organ development and increasing susceptibility to long-term disease^23^. Such redox disturbances may further impair nutrient transport pathways, potentially disrupting the balance of amino acids and lipid metabolism^24,25^.

Metabolomics refers to the systematic identification and quantitation of a wide range of small-molecule weight metabolites in a biological sample, providing a powerful approach to assess small-molecule biochemistry. Furthermore, it offers a comprehensive view of metabolic perturbations caused by complex external exposures, such as tobacco smoking^25^. Previous studies have demonstrated that maternal smoking is associated with alterations in the circulating metabolome, including metabolites associated with endocrine disruption and increased oxidative stress^26–28^.

However, currently, no untargeted (global) metabolomics studies have examined smoking-induced differences in the placenta. Such a study could improve our understanding of how smoking and nicotine exposure affect metabolic processes in the human placenta and the developing fetus. Therefore, in this nested case-control study, we aim to investigate tobacco-induced changes in the placental metabolome using a non-targeted metabolomics approach by analyzing placental samples from women included in our previous Kuopio Birth Cohort (KuBiCo) study^26^.

## Methods

### Placental samples

This study used placental samples from the KuBiCo. KuBiCo is a joint research effort between the University of Eastern Finland (UEF), Kuopio University Hospital (KUH) and Finnish Institute for Health and Welfare (THL)^29^ which was approved by the Research Ethics Committee, Hospital District of Central Finland on 15.11.2011. All participants have given their written electronic consent. The study was conducted in accordance with the Basic & Clinical Pharmacology & Toxicology policy for experimental and clinical studies^30^. The reporting of this study has been done in accordance with the STROBE criteria.

KuBiCo maintains a database of women from the Northern Savo region in Finland who are expected to give birth at KUH. Information on pregnancy and gestational nicotine exposure was gathered from regular maternity clinic visits and via an electronic questionnaire, where they were archived in a database called PikkuHaikara. We also utilized the following descriptive data obtained from the KUH birth registry: maternal age, height, weight, body mass index (BMI in kg/m²), parity, gravidity, length of hospital stay post-delivery, and relationship status.

In the KuBiCo, approximately 1500 placental samples were routinely collected between the years 2012-2020^29^. Here we analyzed placental samples from women whose first-trimester plasma samples we had examined earlier in our previous study^26^, with records of all the background characteristics listed in Table 1. In brief, we utilized samples from women with a parity of six or less, the preference was to use in term placental samples from their first pregnancy. Tobacco smoking was defined as the use of any tobacco product after conception, regardless of the number of cigarettes smoked per day. Based on self-reported data and biochemical confirmation with nicotine metabolite cotinine^26^, participants were categorized into three groups. The control group (n=23) included women who reported no tobacco use during pregnancy and had negative nicotine levels in both first trimester plasma and at term placental samples collected after birth. The early exposure group (n=19) consisted of women with detectable nicotine and/or cotinine levels in first-trimester plasma samples but negative levels in placental samples, indicating early but discontinued use. The continuous exposure group (n=13) included women with positive nicotine and/or cotinine levels in both first trimester plasma and at term placental samples, suggesting ongoing tobacco use throughout pregnancy. Additionally, for those pregnancies where a whole blood sample collected during labor was available (n = 45), we screened for alcohol use using sensitive phosphatidyl ethanol (PEth) measurement^31^, and for drug use using quantitative ultra-high performance liquid chromatography triple quadrupole tandem mass spectrometric (UHPLC-MS/MS) screening covering more than 150 of the most common illicit drugs, designer drugs, and psychoactive pharmaceuticals^32^. Both analyses were performed at the Forensic Chemistry Unit of the Finnish Institute for Health and Welfare (THL) in Helsinki, Finland, and are within the scope of accreditation according to ISO/IEC 17025.

**Table 1:** Background characteristics of the pregnant women.

|  | Control (n=23) |  | Early Exposure (n=19) |  | Continuous exposure (n=13) |  | p value |
| --- | --- | --- | --- | --- | --- | --- | --- |
| Maternal characteristics |  |  |  |  |  |  |  |
| Maternal age (years, mean; ±SD) | 29.04 | ±4.22 | 28.68 | ±7.57 | 26.08 | ±5.94 | 0.335<sup>a</sup> |
| Maternal height (cm, mean; ±SD) | 164.17 | ±5.42 | 164.79 | ±6.54 | 166.54 | ±5.67 | 0.511<sup>a</sup> |
| Maternal weight (kg mean; ±SD) | 61.18 | ±11.16 | 65.11 | ±10.91 | 71.38 | ±11.79 | 0.042<sup>a</sup> |
| Maternal BMI kg/m2 (mean; ±SD) | 22.68 | ±4.06 | 23.91 | ±3.72 | 25.66 | ±3.55 | 0.094<sup>a</sup> |
| Parity (N, mean; ±SD) | 1.39 | ±1.90 | 1.47 | ±2.70 | 1.85 | ±2.03 | 0.836<sup>a</sup> |
| Gravidity (N, mean; ±SD) | 2.78 | ±2.32 | 2.95 | ±2.93 | 4.00 | ±3.16 | 0.422<sup>a</sup> |
| Duration of pregnancy at birth (days, mean; ±SD) | 279.30 | ±8.19 | 277.16 | ±11.39 | 282.00 | ±4.97 | 0.325<sup>a</sup> |
| Pregnant women in committed relationships (%) | 100 | ±0.00 | 94.7 | ±0.23 | 69.2 | 0.48 | 0.007<sup>b</sup> |
| Cigarettes/day before pregnancy (mean; ±SD) | 0.00 | 0.00 | 3.33 | ±8.40 | 4.62 | ±5.94 | 0.066<sup>a</sup> |
| Cigarettes/day during pregnancy (mean; ±SD) | 0.00 | 0.00 | 0.89 | ±2.56 | 1.44 | ±2.40 | 0.134<sup>a</sup> |
| Early neonatal characteristics |  |  |  |  |  |  |  |
| Birth weight (g, mean ±SD) | 3534.57 | ±455.88 | 3527.63 | ±365.61 | 3711.15 | ±352.55 | 0.380<sup>a</sup> |
| Head circumference (cm, mean; ±SD) | 35.14 | ±1.07 | 34.72 | ±1.77 | 35.45 | ±1.11 | 0.330<sup>a</sup> |
| Placental weight (grams, mean; ±SD) | 591.09 | ±123.25 | 595.63 | ±72.89 | 622.31 | ±109.18 | 0.677<sup>a</sup> |
| APGAR scores 1 min (mean ±SD) | 8.96 | ±0.37 | 9.16 | ±0.37 | 9.08 | ±0.28 | 0.184<sup>a</sup> |
| APGAR scores 5 min (mean; ±SD) | 9.17 | ±0.39 | 9.26 | ±0.45 | 9.15 | ±0.38 | 0.702<sup>a</sup> |
| Hospital stay after birth, days | 2.96 | ±1.11 | 3.26 | ±2.10 | 3.00 | ±0.58 | 0.779<sup>a</sup> |
| Postnatal antibiotics (n, %) | 4.3 | ±20.0 | 10.5 | ±31.5 | 0.00 | 0.00 | 0.431<sup>b</sup> |
| NICU admission (n, %) | 8.7 | ±28.8 | 15.8 | ±37.5 | 0 | 0 | 0.324<sup>b</sup> |
| Phototherapy (%) | 0.00 | 0.00 | 5.3 | ±22.9 | 0.00 | 0.00 | 0.395<sup>b</sup> |

Legend: <sup>a</sup>ANOVA test; <sup>b</sup> Chi<sup>2</sup> test.

Placental samples were collected immediately after delivery. Twenty tissue samples, each measuring 2×2 cm, were systematically obtained from the central region of the basal plate and intervillous space, excluding the chorionic plate. These samples were divided into four separate tubes, frozen in liquid nitrogen, and stored at -80°C until the metabolomics analysis^33^. The order of the samples in the metabolomics analysis was randomized using a random number generation algorithm.

### Non-targeted LC-MS based metabolite profiling

Metabolomics analysis was conducted following previously published methods with slight modifications^34^. Approximately 200 µg of placental tissue was placed into homogenizing tubes containing beads. An 80% methanol (MeOH) solution, equal to three times the tissue weight, was added. Homogenization was performed using the Heart Program© (Omni Bead Ruptor) for one cycle at 6 m/s for 30 seconds, followed by sonication for 1 minute at room temperature. Samples were then shaken at room temperature for 5 minutes at speed 10 and centrifuged at 16,100 rcf for 10 minutes at +4°C (Eppendorf Centrifuge 5804R).

The supernatant was diluted 1:1 with 80% MeOH and filtered through a Captiva ND© filter plate by centrifugation (Megafuge 1.0RS, 2,800 rpm, 10 minutes). For quality control, 15 µL from each sample was pooled, vortexed for 10 seconds, and filtered in the same manner. A blank consisting of 80% MeOH was processed identically. All samples were refiltered through the ND filter plate by centrifugation at 700 rcf (1920 rpm) for 10 minutes at +4°C.

Metabolomics analysis was carried out using ultrahigh-performance liquid chromatography (UPLC) with a Thermo Q Exactive Hybrid quadrupole-orbitrap mass spectrometer (MS). Samples were analyzed by two chromatographic techniques—reversed-phase (RP) and hydrophilic interaction liquid chromatography (HILIC). RP chromatography utilized a Zorbax Eclipse XDB-C18 column, with mobile phase A consisting of water with 0.1% formic acid (HCOOH) and mobile phase B composed of methanol with 0.1% formic acid, following a 16.5-minute gradient at a flow rate of 0.4 mL/min. The HILIC setup employed an Acquity BEH amide column, where mobile phase A consisted of 50% acetonitrile with 20 mM ammonium formate buffer, and mobile phase B contained 90% acetonitrile with 20 mM ammonium formate buffer, on a 12.5-minute gradient at a flow rate of 0.6 mL/min.

Electrospray ionization (ESI) was used in both positive and negative modes. The ESI parameters included a spray voltage of 3.5 kV for positive mode and 3.0 kV for negative mode, with a maximum spray current of 100. The flow rates were set at 40 for sheath gas, 10 for auxiliary gas, and 2 for spare gas (measured in arbitrary units for the ion source). Additional settings included an S-lens RF level of 50 V, and capillary and probe heater temperatures maintained at 300°C. The full scan range covered m/z 60 to 700 in HILIC mode and m/z 120 to 1200 in RP mode, with a resolution of 70,000 (m/Δm, full width at half maximum at 200 u), and automated injection time and gain control targeted at 1,000,000 ions.

For tandem mass spectrometry (MS/MS), three peaks were selected for fragmentation using an apex trigger ranging from 0.2 to 3 seconds, with a dynamic exclusion of 15 seconds. Normalized collision energy values of 20, 30, and 40 were applied, with a mass resolution of 17,500 (m/Δm, full width at half maximum at 200 u), an automated gain target of 50,000, and an isolation window of 1.5 m/z.

Data processing was performed using MS-DIAL software version 4.90, with peak picking, alignment, and metabolite identification. Data collection settings included an MS1 tolerance of 0.005 and an MS2 tolerance of 0.025, with a retention time range between 0.5 and 100 minutes. The mass range for MS and MS/MS analyses extended from 50 Da to 2000 Da. Peak detection parameters were configured with a minimum peak height of 300,000, an amplitude smoothing level of 3 scans, a minimum peak width of 8 scans, and a mass slice width of 0.1 Da. Deconvolution settings included a sigma window of 0.5 and an amplitude cut-off of 0.

Metabolite identification was carried out using the University of Eastern Finland’s in-house database, with MS1 and MS2 mass tolerances set at 0.008 Da and 0.025 Da, respectively. Automatic identifications were reviewed and manually corrected when necessary. Adduct ion settings for positive ionization included [M+H]+, [M+NH4]+, [M+Na]+, [M+K]+, [M+Li]+, [M+ACN+H]+, [M+H-H2O]+, [M+H-2H2O]+, and [2M+H]+, while those for negative ionization comprised [M-H]-, [M-H2O-H]-, [M+Na-2H]-, [M+Cl]-, [M+FA-H]-, [M+Hac-H]-, and [2M-H].

Alignment parameters were defined with a retention time tolerance of 0.05 minutes and an MS1 tolerance of 0.015. Molecular features detected in at least 30% of samples within a group were included, with a gap-filling strategy applied when necessary.

Data preprocessing, including quality assessment, drift and batch effect corrections, and imputation of missing values, was conducted using the R-package notame^30^. Metabolite identification was conducted by focusing on molecular features with p-values less than 0.05. These features were classified into four levels based on established community guidelines^35^. Level 1 identification included compounds that were confirmed through matching exact mass, isotope pattern, retention time, and MS2 fragmentation with chemical standards from the in-house library. Level 2 identification encompassed putatively annotated compounds, identified by comparing exact mass and MS2 spectra against public databases. Level 3 identification involved compounds characterized at the chemical group level but without precise compound identification, while Level 4 consisted of unidentified compounds. The identified metabolites were grouped according to the Human Metabolome Database (HMDB)^36^.

### Statistical analysis

Welch’s t-test was used for continuous variables and Chi2 test for categorical variables in the background characteristics. For the metabolomics data, effect sizes were calculated using Cohen’s d method and p-values were calculated with Welch’s t-test. Because molecular features in a non-targeted metabolomics data are not independent, the number of principal components needed to explain 95% of the variance in the metabolomics data in a principal component analysis (PCA) was used to account for multiple testing using the Bonferroni method. SIMCA (version 17, Umetrics, Sartorius Stedim Data Analytics AB) was used for the PCA and molecular features were scaled and centered by z-score normalization and dataset was split into 7 subsets for cross-validation. Here, 47 principal components were needed to explain 95% of the variation in the metabolomics data and therefore the α level was adjusted to 0.001. Findings with p-values between 0.05 and 0.001 were considered to be trends. Comparisons used were early exposure vs controls, continuous exposure vs controls, and both early and continuous exposure vs controls. R-packages notame, ComplexHeatmap 2.26.0 ja circlize 0.4.17 were used for statistics and making figures.

## Results

Table 1 summarizes the characteristics of the pregnant women and their newborns. Maternal characteristics showed that women in the early and continuous exposure groups had significantly higher body weight compared to the control group. Additionally, a significantly higher percentage of women in the control group were in a relationship compared to the exposed groups. No other maternal characteristics showed statistically significant differences. Similarly, there were no significant differences in any of the newborn characteristics across the three groups. Moreover, all whole blood samples screened for alcohol exposure with PEth were negative (PEth < 2ng/mL), indicating no recent alcohol exposure in the subjects. No illicit or designer drugs were detected in any of the samples. Pain medications paracetamol was detected in 30% and oxycodone in 2% of the whole-blood samples collected during labor.

In the metabolomics analysis, there were differences between the controls and cases (early exposure and continuous exposure groups). These are shown in Figure 1 and Supplementary Table S1. However, most of changes in identified metabolites had a p-value between 0.05 and 0.001 and should therefore be considered as trends that need to be validated in future research.

**Figure 1:**
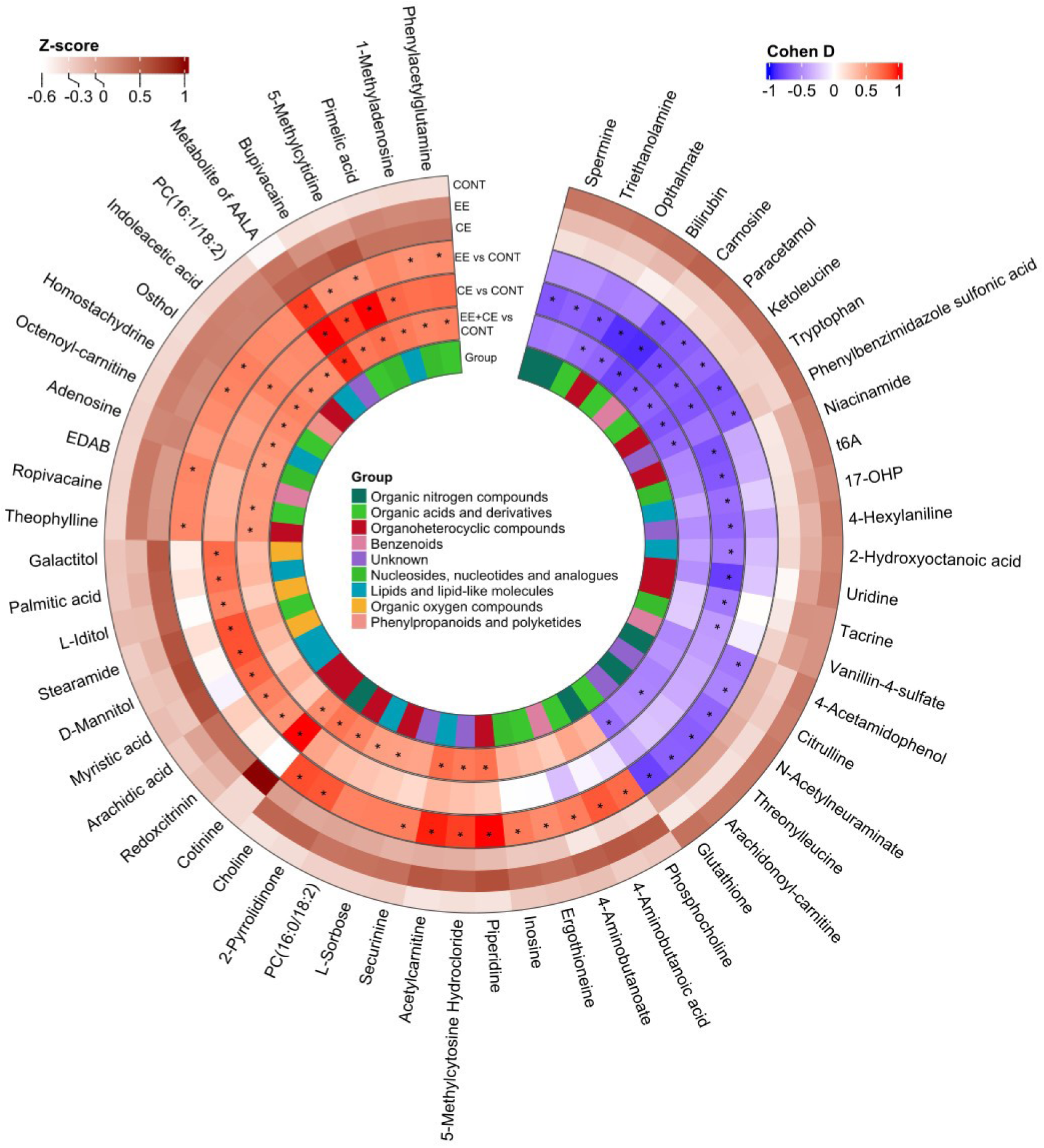
Circular heatmap of placenta metabolites. Identified metabolites with p-value below 0.05 in any comparison between the study groups are shown. CONT = Control group, EE = Early Exposure group, i.e., placental samples from pregnancies where the woman positive cotinine levels in 1^st^ trimester serum samples but no cotinine in the placental sample, and CE = Continuous Exposure group, i.e., placental samples from pregnancies where both the 1^st^ trimester and placental sample had high cotinine levels. Z-scores are shown for each group, and Cohen’s d effect sizes are shown for each group comparison. *, p-value < 0.05. Metabolite of AALA, metabolite of amino acid local anesthetic; EDAB, ethyl-4-dimethylaminobenzoate; t⁶A, N⁶ threonylcarbamoyladenosine, 17-OHP, 17α-Hydroxyprogesterone.

As expected from the study design, cotinine levels were significantly higher in the continuously exposed group compared to control group (Figure 1, Supplementary Table S1). Several measured molecular features, such as metabolite of aminoamide local anesthetic, bupivacaine, ropivacaine and 5-methylcytidine levels we high in both early exposure and continuously exposed groups when compared to controls (Figure 1, Supplementary Table S1). Furthermore, also some metabolites, like carnosine and tryptophan levels were low in both exposed groups when compared to controls (Figure 1, Supplementary Table S1). However, there were also group specific changes. For example, the continuous exposure group had low levels of uridine and bilirubin when compared to the controls (Figure 1, Supplementary Table S1). Whereas, in the early exposure group, we observed low levels oxidized glutathione and high ergothioneine levels when compared to the controls (Figure 1, Supplementary Table S1).

## Discussion

The results of this exploratory study showed differences in the placental metabolite profiles between non-smoking pregnant women and those who either quit smoking during pregnancy (early exposure) or continue to smoke throughout the pregnancy (continuous exposure). Main results indicate alterations in placental enzyme activity, xenobiotic metabolism, as well as amino acid metabolism and oxidative stress. These observations align with previous studies in pregnant women, as well as with targeted analyses examining tobacco’s effects on placental metabolism^28,37^. For most metabolites, the differences observed in both early exposure and continuous exposure groups were in the same direction when compared to the controls, although the effect size varied. This pattern suggests that many of the tobacco smoking associated effects to placental metabolome are outlasting the acute effects of nicotine and other chemicals in tobacco.

Nicotine metabolite cotinine, as expected by the study design, was significantly higher in the placental samples from pregnant women who continue to smoke throughout the pregnancy. This confirms appropriate group classification and validates this metabolomics approach. Also, this marks placental cotinine as a good biomarker for continuous smoking exposure^38,39^.

Furthermore, homostachydrine (pipecolic acid betaine) levels were high in the early exposure group. Homostachydrine is found in many plants, but especially in coffee^40^,indicating higher coffee intake those who quitted smoking during pregnancy. This is in line with other findings, like high theophylline (a xanthine found in tea and coffee) levels in the early exposure group

An unknown metabolite of aminoamide local anesthetic was significantly more abundant in the placental samples of both early exposure and continuous exposure groups when compared to the controls. Additionally, levels of ropivacaine and bupivacaine were higher in both exposure groups. Ropivacaine and bupivacaine are commonly used local anesthetics during labor, which explains their presence in placental samples^41^. The elevated placental levels of the metabolite of aminoamide local anesthetic may be linked to the induction of CYP enzymes, which are upregulated by the exposure to polycyclic aromatic hydrocarbons present in tobacco^42^. This enzymatic induction enhances the metabolism of numerous xenobiotics, including anesthetics, leading to altered pharmacokinetics^43^. Moreover, in line with previous literature, smokers typically require higher doses of anesthesia and analgesics due to these metabolic adaptations^44^. Therefore, the higher abundance of the aminoamide local anesthetic metabolite in the exposed groups may reflect both the accelerated metabolism of anesthetics in smokers, and an increased clinical demand for the anesthetic agents.

We found that pregnant smokers had alterations in placental levels of some dipeptides and amino acids, such as carnosine and tryptophan. Previous studies have demonstrated that smoking may modify nutrient metabolism, leading to altered amino acid profiles in smokers^45,46^. Reduced nutrient intake and changes in hepatic metabolism could partially explain the decreased abundance. These findings expand on earlier researches^47,48^, by suggesting that chronic smoking exposure not only alters appetite but also disrupts metabolic recovery, influencing the ion abundance of several metabolites in biological tissues like placenta compared to nonsmoking controls. Poor nutrient intake is also supported by, e.g., the low niacinamide (B3) levels seen in the continuous smoking group.

In addition, we observed significant alterations in cellular antioxidants and detoxifiers, especially in the early exposure group, such as low levels of oxidized glutathione and high ergothioneine levels, when compared to the controls. This indicates reduced oxidative stress in the early exposure group, which is a good sign, since antioxidants, like ergothioneine, are known to provide metabolic homeostasis by neutralizing damaging ROS^49^ and reduced antioxidant availability increases the risk of complications for both the mother and fetus, as it impairs organogenesis, causes placental dysfunction and changes postnatal health^50^. Prolonged smoking or nicotine exposure may chronically activate oxidative stress responsive pathways, stimulating upregulation of antioxidant and detoxification systems such as glutathione synthesis. The effects seen in the early exposure group could be due to upregulated system not yet down regulated even though the cause of the oxidate stress, here smoking, is reduced or stopped. Overall, these results indicate that quitting smoking during pregnancy does seem to reduce oxidative stress in the placenta.

This study has limitations. Firstly, tobacco smoking was self-reported by the pregnant women. It is well established that individuals often underreport their substance use in self-reported surveys^51^. However, this bias was minimized by measuring nicotine and cotinine levels in plasma samples during the first trimester and in placental samples at the time of birth. Furthermore, combined tobacco and alcohol use is common^52^, and alcohol during pregnancy is known to alter metabolite profiles^53^. However, here we were able to confirm that there had not been any recent alcohol exposure using sensitive PEth analysis^31^, indicating that the effects observed in this study are associated with tobacco exposure rather than alcohol exposure. Additionally, this was an explorative study with limited sample size. The study design required pairing placental samples with previously collected first-trimester plasma samples, but not all women had placental material available for analysis. Subdividing participants into early and continuous exposure groups further reduced group sizes, which limited the statistical power to detect differences in certain metabolites. Moreover, the study population was primarily from the Northern Savo region of Finland, participants likely shared similar lifestyle habits, including dietary patterns. Furthermore, the typical effects that are expected to be seen when comparing smoking and nonsmoking pregnant groups, like reduced newborn weight and head circumference, were not observed on the group level comparisons. This suggests selection bias, as it is likely that women who are heavy users of tobacco are less willing to have their placentas collected for analysis. Finally, for women in the early exposure group, the exact timing of smoking cessation during pregnancy was not available, which introduces uncertainty into exposure classification. Overall, to confirm these findings, it is important to replicate the study in larger cohorts with different population backgrounds.

In conclusion, maternal smoking during pregnancy induces alterations in placental metabolism, particularly in amino acid metabolism, oxidative stress defenses, and xenobiotic metabolism. Most observed alterations were similar between early exposure and continuous exposure groups, although differences in the effect sizes were seen, indicating that tobacco exposure leads to long lasting changes in the placental metabolome. These findings emphasize the role of placenta in the effects of tobacco during pregnancy. Further studies are needed in different populations and larger sample sizes to broaden the applicability of these results.

## Data Availability

All data produced in the present study are available upon reasonable request to the authors.

## Acknowledgements

This work was supported by the Finnish Ministry of Social Affairs and Health for the VAURAS project, and the Finnish Cultural Foundation. This paper belongs to the studies carried out by the Kuopio Birth Cohort consortium (https://www.KuBiCo.fi) and we thank our colleagues who are responsible for the design and conduct of the KuBiCo. We want to thank Miia Reponen and Juulia Kuparinen for technical assistance with the mass spectrometry analyses and the staff of the Department of Obstetrics and Gynecology in Kuopio University Hospital. The authors also want to thank Biocenter Finland and Biocenter Kuopio for supporting the core LC-MS laboratory facility. We would also like to thank Ms Sanna Kyllönen of the Finnish Institute for Health and Welfare (THL) and Mr Antti Jylhä (THL) for their contribution to the laboratory analyses.

## Conflict of interest

O.K. is a co-founder of Afekta Technologies Ltd., a company providing metabolomics analysis services. W.M, R.H, T.G., M.L, L.K-N, J.R declare no conflicts of interest.

## Data Sharing Statement

The data that support the findings of this study are available on request from the corresponding author, O.K. The study plan approved by the ethical committee and the participant consent terms preclude public sharing of these sensitive data, even in anonymized form.

## Notes

### Funding Statement

This study was funded by the Finnish Ministry of Social Affairs and Health and the Finnish Cultural Foundation.

### Author Declarations

Research Ethics Committee of the Hospital District of Central Finland gave ethical approval for this work.

